# Horizon Sham Adaptor: Unlocking the Potential of Dual-Functionality of Single TMS Coil for Real and Sham Stimulations

**DOI:** 10.1101/2023.10.17.23297143

**Authors:** Majid Memarian Sorkhabi

## Abstract

This paper introduces a new figure-of-eight TMS coil design capable of delivering both real and sham TMS stimulation using a single coil. The design, emitted clicking noise, and induced tactile sensation provide no discernible indication of whether it is in the real or sham state, making it suitable for double-blind studies. This coil family comprises the E-z Cool Coil and Air Film Coil (AFC) and is compatible with Magstim Horizon stimulators. This single-coil TMS setup enables the administration of either real or sham protocols without requiring coil replacement or physical repositioning. The transition between these states is achieved simply by replacing the adaptor between the coil connector and the TMS device.

## I. Introduction

Transcranial magnetic stimulation (TMS) is a highly versatile and attractive non-invasive technique that finds applications across both the realms of neuroscientific research and clinical diagnosis and therapy. It operates on the core principles of magnetic induction to modulate the central nervous system. This method involves the application of a brief electric current to a treatment coil, which then generates a rapidly changing magnetic field. This dynamic magnetic field induces a voltage within the brain tissue located underneath the coil, thereby enabling precise and controlled stimulation. Beyond its significance in research, TMS has gained recognition for its therapeutic potential, particularly in addressing conditions such as major depressive disorder (MDD), obsessive-compulsive disorder (OCD), and anxious depression. Its non-invasive nature and ability to target specific brain regions make TMS a promising intervention, offering new avenues for treatment and enhancing our understanding of the intricacies of the human brain.

In psychiatric clinical studies, particularly when evaluating the efficacy of treatments like TMS, controlled studies provide a critical framework for investigating the true impact of psychiatric interventions. Open-label studies lack controls, making them susceptible to biases and placebo effects, while single-blind studies introduce the potential for investigator bias. In contrast, double-blind, controlled studies set the gold standard by concealing both the treatment assignment and its administration, thus minimizing biases and external influences. This rigorous methodology enhances the reliability of findings and the ability to establish causal relationships between TMS and psychiatric outcomes, essential given the complexity of mental health conditions. Controlled studies play a pivotal role in advancing evidence-based psychiatric practices, ensuring that TMS and similar interventions are not only safe but also effective in improving the well-being of individuals struggling with psychiatric disorders.

Addressing the potential confounding effects of ancillary factors, such as clicking sounds and scalp sensations, is crucial in TMS studies to prevent disruptions in task performance and study participant bias. With the substantial placebo effects in clinical research, especially in studies involving medical devices with substantial patient-investigator interaction, distinguishing the genuine effects of brain stimulation from these artifacts is essential. To achieve this, TMS experimenters often employ control conditions where they administer either sham (placebo) stimulation or real (standard or active) stimulation. These complementary approaches aim to ensure that differences between experimental and control conditions solely arise from the method of brain stimulation, while any auditory or tactile artifacts closely mimic real stimulation. This strategy not only allows for the separation of these artifacts but also aids in demonstrating the specificity of the targeted brain region. However, creating a valid sham condition can be challenging, as it requires replicating the sensory side effects of TMS in the absence of an actual magnetic field, including rhythmic scalp muscle stimulation and clicking sounds produced during coil discharge, which may necessitate specialized equipment.

### II. Literature review

Currently, many research methods fall short of achieving the ideal double-blind interleaving of stimulus conditions and introduce their own set of experimental challenges. Various techniques have been employed to mitigate the impact of TMS field redirection. One approach involves tilting the coil away from the scalp, ranging from 45° to 90° away from the target brain area [1], or stimulating a control brain site like the vertex [2] [3]. However, these methods fail to fully control for physical sensations, mainly limited to the study participant’s perception of scalp contact with the coil. Additionally, in certain cases, tilted coils still manage to deliver effective levels of stimulation. When stimulating control brain sites, the potential confounding effects remain largely unknown. Moreover, the necessary coil movements to switch between conditions pose difficulties in maintaining stable stimulation sites.

In the context of zero-field stimulation, which is another approach for sham stimulation, two primary categories of techniques have been utilized: simulating peripheral sensations while the coil remains inactive and utilizing dedicated sham coils. In the first category, some researchers replicated the coil noise by discharging an additional TMS unit with its coil positioned nearby. Others combined this with electrical stimulation to induce scalp sensations [4]. In some studies, disposable pad electrodes (referred to as “sham pads”) were used to deliver synchronous direct current stimulation, mimicking the tactile sensations of active stimulation. However, it’s worth noting that this method is not compatible with electroencephalogram (EEG) equipment [5].

Utilizing dedicated sham coils presents an additional drawback in that they must be physically exchanged with the standard coil. This introduces the challenges related to maintaining consistent localization, repositioning neuro-navigation trackers and tools, and compounds the inconvenience with the need to switch heavy cables and perform (re-)clamping of the coil holder. Consequently, not only does this make the process of interleaving conditions highly challenging, but it also increases the study participant’s awareness of the experimental manipulation.

## III. Method

The objective of designing the new Magstim sham coils is to create and validate a method for delivering both real and sham TMS conditions using a single coil. Basically, the new design features a figure-of-eight coil with loops of coils in each of its two wings, each driven independently. Different adaptors enable the administration of either real or sham stimuli without the need to switch or reposition the coil, as illustrated in Figure 1. Real stimuli are provided when the current direction in both figure-of-eight loops agrees with the standard treatment coils and sham stimuli are provided when the direction of the current in one of the two loops is reversed (Figure 1(a) and Figure 1(b)).

**Figure 1.**
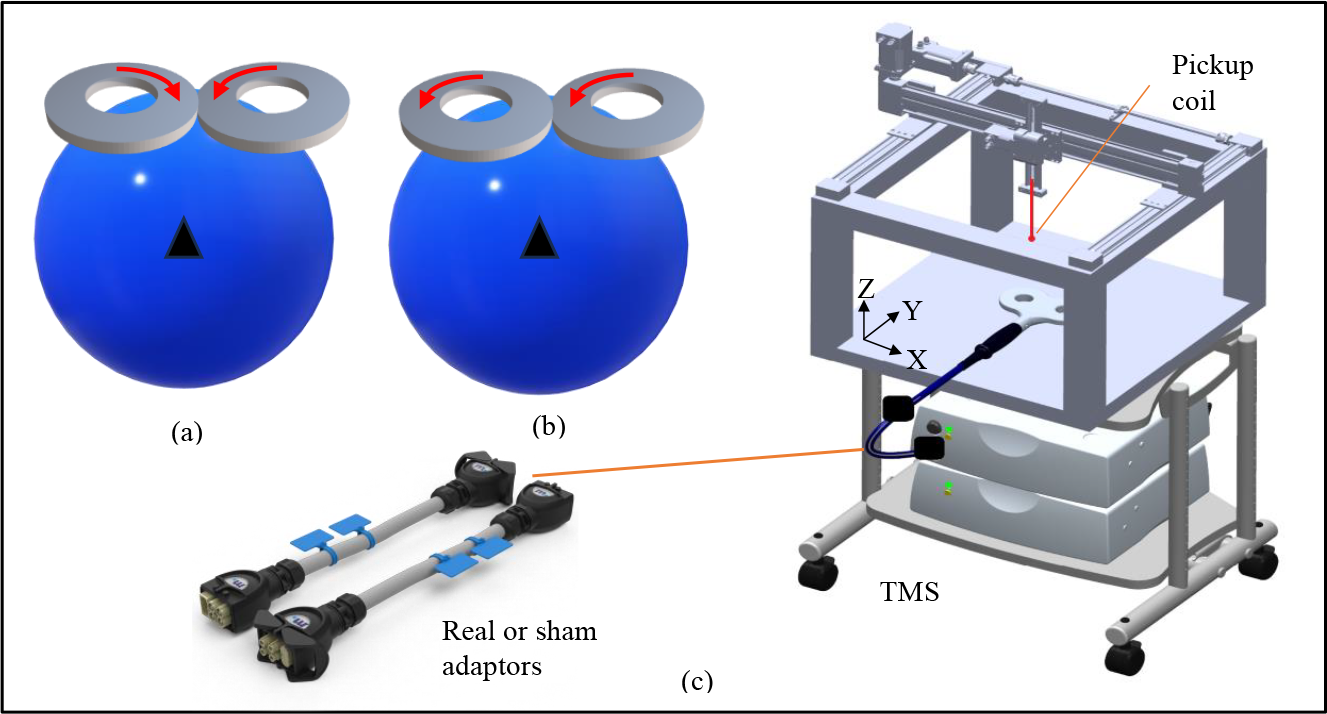
The concept of a single coil for real and sham conditions with two adaptors (a) Real state of the coil, (b) Sham state of the coil. The electric current’s direction is indicated by the red arrows on the coils. (c) E-field mapping setup for the measurements, using the 3D pickup coil. Peak E-field spatial maps were acquired. Data was collected with 5 mm steps from X = -10 cm to X = +10 cm, Y = -5 cm to Y = +5 cm, and Z = 0 to Z = 4.5 cm, within the region of interest.

We conducted both mathematical modeling and practical measurements employing a pickup coil to assess the physical and spatial attributes of the stimuli, including electromotive force (EMF). We performed electromagnetic simulations using the COMSOL software, using the finite element method (FEM) and the AC/DC module. For these simulations, a spherical head model with a diameter of 17 cm was employed to determine the induced electric field (E-field). The head tissue was assumed to be uniform, with an electrical conductivity of σ = 0.33 S/m [6]. During the FEM simulation, both the treatment coil and head were positioned within an air environment, similar to [7].

For the EMF measurement, a custom made 3D circular pickup coil was used with diameter of 5 mm. Measurements were done with a custom designed robotic tool to move the pickup coil all around the treatment coil in 5 mm steps and 3 directions, with 10 µm accuracy, as shown in Figure 1. The pick-up coil EMF was recorded with a calibrated digital oscilloscope (Tektronix MDO34, Tektronix, OR USA). We used the same pickup coil for all measurements to eliminate any potential relative differences between them. This verification method is similar to [8].

## IV. Results

### IV. I Modeling results

In the real coil state, the peak E-field is induced in the overlap of the two wings, as shown in Figure 2 (a). In the case of sham stimulation (Figure 2(b)), it is evident that within the central region and beneath the overlap of the two wings, the E-fields generated by the two coils mutually attenuate, leading to a lack of effective stimulation in the deeper brain regions. However, in areas located below the coils, the potential for stimulation becomes evident. This off-target stimulation on the scalp can evoke a sensation of stimulation in the study participant, potentially augmenting the realism of the placebo effect.

**Figure 2.**
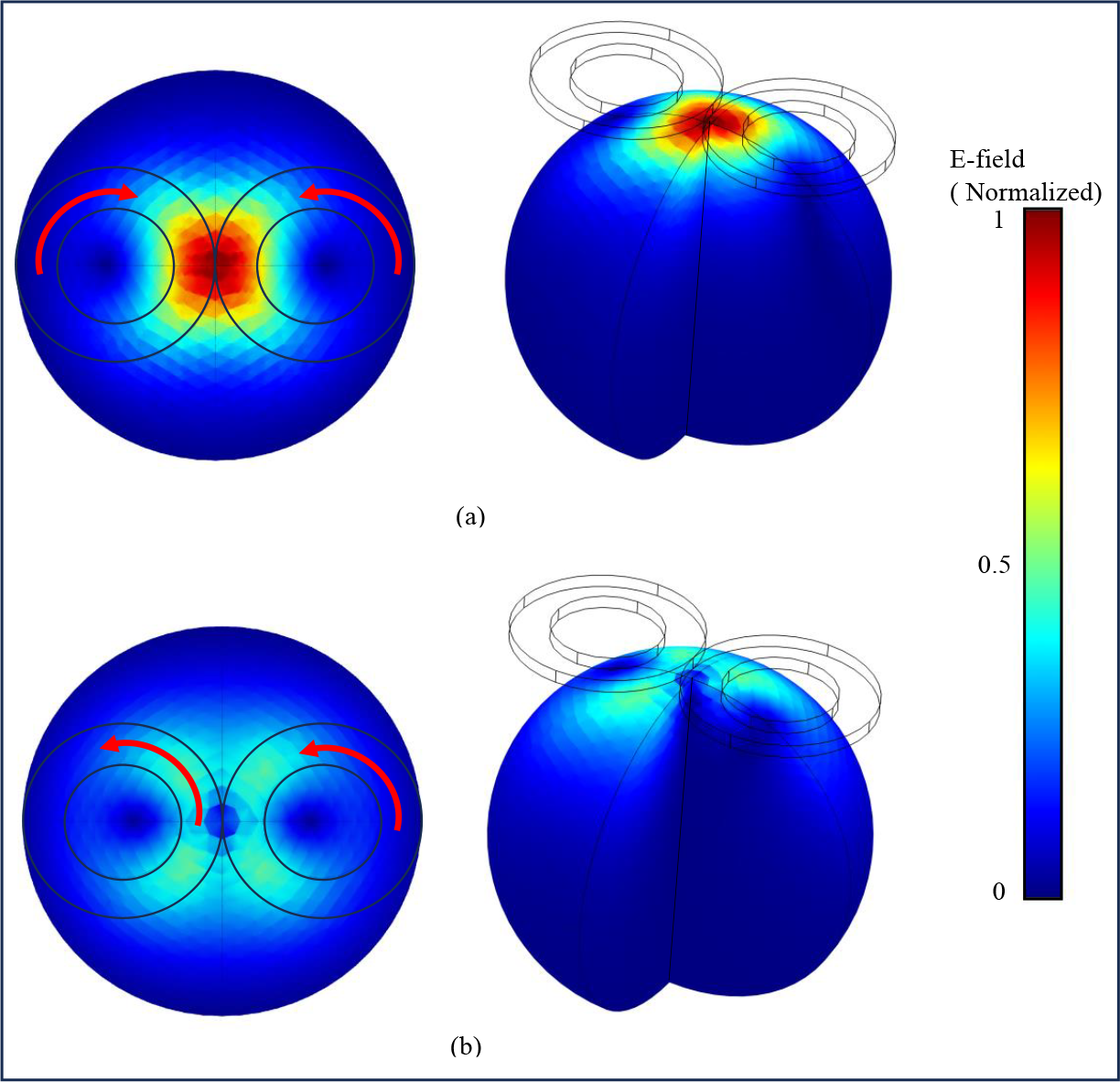
Computational modeling results illustrating the spatial distribution of induced E-field norm for (a) the real state and (b) the sham state of the coil. The red arrows on the coils indicate the direction of the electric current.

### IV. II Measurement results

The measurement findings closely correspond with the modeling outcomes, highlighting a significant alignment between the two datasets. The measurement results are shown in Figure 3 and Figure 4, as 3D views of the region of interest and coronal plane cross-sections of the head, respectively. The outcomes reveal that, in certain regions, the induced E-field on the scalp and up to a depth of 1 cm inside the head exhibits similarity between the real and sham coils, while in some areas, the sham coil produces different field strength. In Figure 4 (c) areas where the E-field of the sham state approximates the field generated by the real coil are identified with diagonal stripes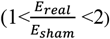. In these regions, it is anticipated that the tactile sensation induced by the sham state will closely resemble that of the real state.

**Figure 3.**
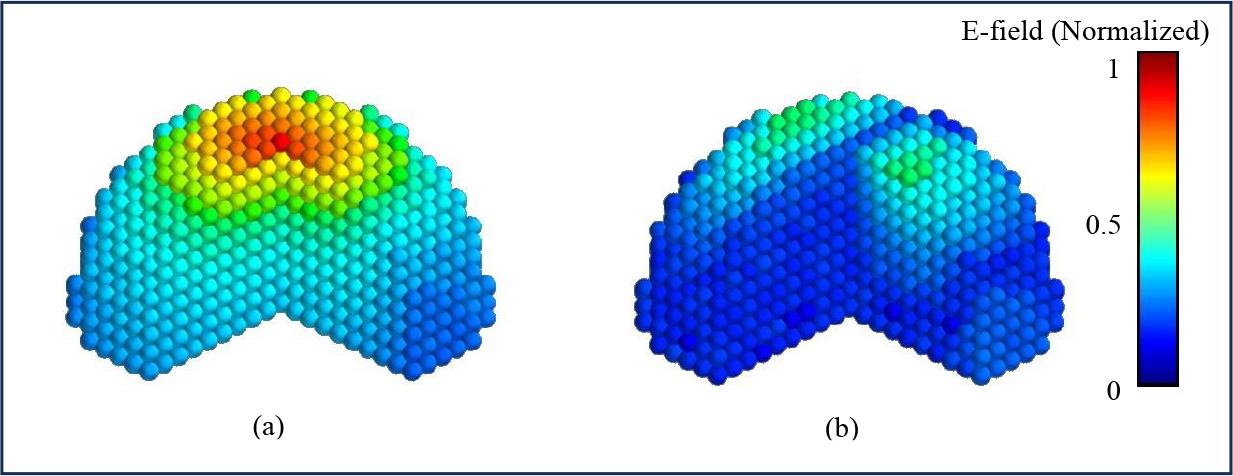
Measurement results depicting the spatial distribution of induced E-field in the region of interest for (a) the real state and (b) the sham state of the coil. Each small sphere shows a 5 mm measurement step.

**Figure 4.**
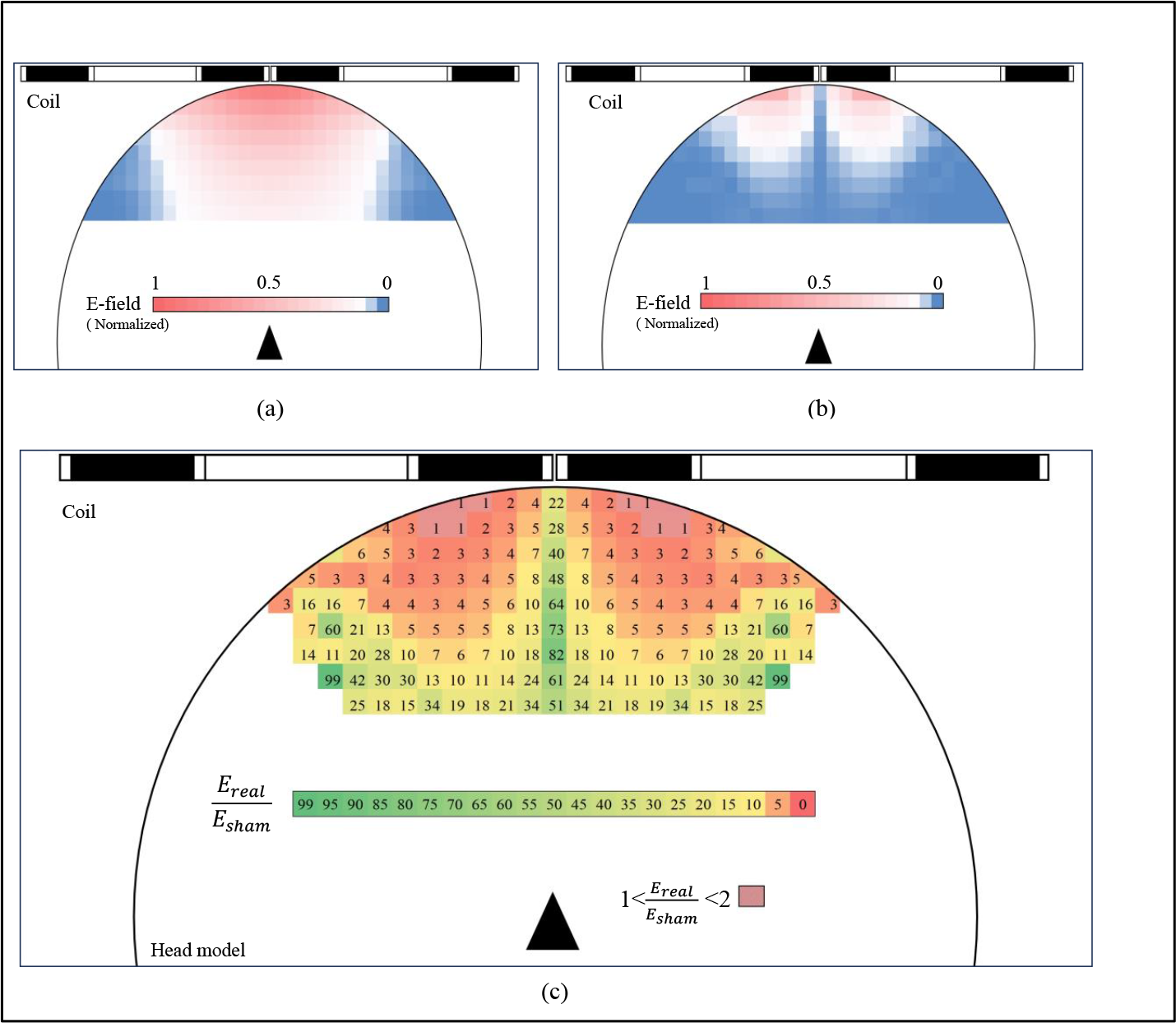
Measurement results for induced E-field within a coronal plane cross-section of the head, with 5 mm intervals between measurements. Each square corresponds to a 5mm x 5mm area of the brain. The induced E-field in (a) the real state and (b) the sham state of the coil. (c) The ratio between E-fields in the real and sham states. Ratios are rounded to the nearest integer.

Conversely, at greater depths, specifically 1.5 cm, 2 cm, 3 cm, and 4 cm beneath the skull’s surface, the induced field by the real state is, on average for the region of interest area, 10, 13, 20, and 25 times higher, respectively, than that produced by the sham state. Consequently, the potential for induced neuromodulation by the sham state in deeper cortical areas is significantly limited.

### IV. III Measurement of Sound Pressure Levels

We conducted measurements to determine the maximum sound pressure levels produced by both the real and sham states of the coil (AFC and E-z Cool coils) at 100% maximum output. These measurements were taken using a sound pressure monitor, with measurements recorded at a distance of 10 cm from the coil surface, and each condition was tested 20 times. To assess any significant differences between the conditions, we performed paired t-tests. Our analysis revealed that there were no statistically significant differences in sound levels between the two coil states (all p-values > 0.4). As an example, in the real state of the AFC coil, the maximum sound pressure level measured 89.2 dBA, whereas in the sham state, it was recorded as 91.1 dBA.

### IV. Measurement of E-field shape

We conducted measurements to characterize the induced E-field shape generated by both the real and sham states of the coil. These measurements were performed using the same pickup coil, at an off-centre point. The results of these measurements are presented in Figure 5.

**Figure 5.**
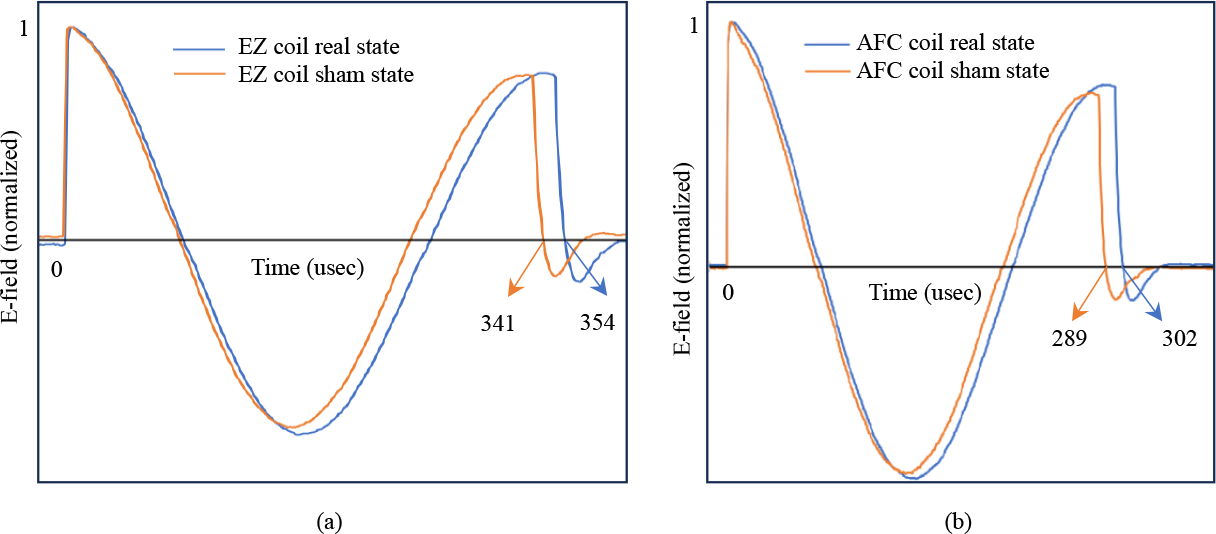
Recorded waveforms from the coils representing real and sham pulse waveforms. (a) Presents normalized E-field waveforms for both Ez in real and sham states. (b) Displays normalized E-field waveforms for AFC in real and sham states. Recording were obtained at a sampling rate of 1 Mega samples per second.

In the case of the Ez coil, the pulse width for the real state was determined to be 354 usec, while for the sham state, measured 341 usec. Similarly, for the AFC coil, the pulse width in the real state measured 302 usec, whereas in the sham state, measured 289 usec. Across both coils and states, the discernible difference in pulse width consistently stays below 13 usec. This observed variation in pulse width for the real and sham states can be primarily attributed to differences in mutual coil induction between the two wings of the figure of eight coil. Consequently, we do not anticipate any recognizable variations in the clicking sounds emitted by the coils for either the real or sham states.

## V. Discussion

This paper introduces a figure-of-eight coil designed for both real and sham stimulation, featuring active cooling. We assessed the spatial and temporal characteristics of these stimulation modalities through computational modelling and E-field measurements. Notably, the profiles of real stimuli closely resembled the E-fields generated by conventional Figure of eight coils. Our findings reveal a significant reduction in E-field amplitude when comparing the sham state to the real state, especially in deeper brain regions.

These novel adaptors and coils empower researchers to manipulate variables, employ placebo controls, and implement randomization, all of which are essential for accurately evaluating the effectiveness of psychiatric interventions. This level of control is particularly critical in psychiatry, given the complex nature of mental health conditions. Controlled studies, by mitigating biases and external influences, enable the establishment of causal relationships between treatments and outcomes. This approach offers a higher degree of certainty in assessing the benefits and limitations of psychiatric interventions [9].

### VI. Recommendations for researchers

1-First determine the motor threshold level or motor hotspot on the participant’s head using the coil directly connected to the TMS device, without any adaptors. In this configuration, the coil state is set to “real.” After successfully locating the motor threshold and securing the coil in place, you can then connect the appropriate adaptor to the coil connector to switch between real and sham protocols as needed.

2-Similar to all double-blind studies, the control of treatment protocols is typically overseen by a third party, often referred to as the “study coordinator” or “blinded investigator”. This individual is responsible for providing the appropriate adaptor to administer either real or sham treatment, following a predetermined randomization code or protocol. While the operator cannot differentiate between the real and sham types based on the appearance of the adaptor or the emitted click noise, they can identify it by reading the part number on the label and cross-referencing it with the user manual. Therefore, it is advisable to cover the adaptor’s label with an appropriate sticker before handing it over to the operator.

### VII. Limitation

1-The sham TMS method introduced in this research, as well as in other related research [6] [10], induces subjective somatic sensations in the scalp and facial muscles during sham stimulation due to off-target stimulation. While this off-target stimulation shares similarities with the real state, it may exhibit higher intensity levels at certain points and can vary from person to person.

2-Horizon Sham Adaptors are not intended for use in investigations or studies involving peripheral stimulation. While this sham state exhibits similar E-field characteristics on the coil surface, the field strength in the depth of 1 cm and below is significantly diminished, especially in the central region of the coil. This depth parameter gains significance when considering the relative thickness of the scalp and skull. However, in the context of peripheral magnetic stimulation, where neuromuscular tissues are in much closer proximity to the skin, the sham state may potentially produce similar stimulation effects on peripheral tissues.

## VIII. Data Availability

All data and modelling files are available upon request. Contact Magstim for access to raw and processed data, including computational and measurement results.

## IX. Disclaimer and Legal Notice

While every effort has been made to ensure the accuracy and reliability of the content, the information presented in this paper is intended for informational purposes only and should not be construed as legal, medical, or professional advice. It is the responsibility of readers to verify the legal and regulatory requirements in their jurisdiction and to comply with all applicable laws and guidelines when implementing the recommendations or strategies discussed in this paper.

## X. Contact Information

For product specific operational training, installation or for further information, please contact your local Magstim® representative directly. The latest safety summary for the devices contained within this operating manual may be viewed via the European Databank on Medical Devices.

Online: www.magstim.com/contact

Global:

The Magstim® Company Ltd. Spring Gardens, Whitland, Carmarthenshire, UK, SA34 0HR

Tel: +44 (0)1994 240798

Servicing Enquiries Tel: +44 (0)1994 242900

Sales Enquiries Tel: +44 (0)1994 241111

**European Authorized Representative**: Technomed Europe Amerikalaan 71, 6199 AE Maastricht-Airport The Netherlands

Tel: +31 43 408 68 68

, website: www.technomed.nl

## References

[1] S. H. Lisanby and et al., “Sham TMS: intracerebral measurement of the induced electrical field and the induction of motor-evoked potentials,” Biological Psychiatry, vol. 49, no. 5, pp. 460–463, 2001.

[2] C. Nettekoven and et al., “Dose-Dependent Effects of Theta Burst rTMS on Cortical Excitability and Resting-State Connectivity of the Human Motor System,” J Neurosci., vol. 34, no. 20, p. 6849–6859, 2014.

[3] K. Wendt and et al., “The effect of pulse shape in theta-burst stimulation: Monophasic vs biphasic TMS,” Brain Stimulation, vol. 16, no. 4, pp. 1178–1185, 2023.

[4] J. J. Borckardt and et al., “Development and evaluation of a portable sham transcranial magnetic stimulation system,” Brain Stimulation, vol. 1, no. 1, pp. 52–59, 2008.

[5] E. J. Cole and et al., “Stanford Neuromodulation Therapy (SNT): A Double-Blind Randomized Controlled Trial,” American Journal of Psychiatry, 2022.

[6] M. Memarian Sorkhabi and T. Denison, “A neurostimulator system for real, sham, and multitarget transcranial magnetic stimulation,” Journal of Neural Engineering,, vol. 19, no. 2, 2022.

[7] M. Memarian Sorkhabi and et al., “Temporally interfering TMS: focal and dynamic stimulation location,” in 42nd Annual International Conference of the IEEE Engineering in Medicine & Biology Society (EMBC), 2020.

[8] L. I. N. de Lara and et al., “A 3-axis coil design for multichannel TMS arrays,” NeuroImage, vol. 224, 2021.

[9] F. Duecker and A. T. Sack, “Rethinking the role of sham TMS,” Frontiers in Psychology, 2015.

[10] F. Hoeft and et al., “Electronically Switchable Sham Transcranial Magnetic Stimulation (TMS) System,” PLoS One, 2008.

